# Understanding Antimicrobial Stewardship in Skilled Nursing Facilities Through a Complex Adaptive Systems Perspective: A Qualitative Study in Southern Arizona

**DOI:** 10.64898/2026.03.23.26349116

**Authors:** Flavia Nakayima Miiro, Theresa A. LeGros, Connor P Kelley, James K. Romine, Katherine D. Ellingson

## Abstract

**Introduction:** Antibiotic use drives antimicrobial resistance, and optimizing prescribing in skilled nursing facilities (SNFs) – which care for medically complex residents in congregate settings characterized by frequent care transitions and diagnostic uncertainty – presents unique challenges. Antimicrobial stewardship (AMS) in SNFs has therefore become a focus of quality improvement efforts by federal and state health agencies. We aimed to identify factors that facilitate and hinder AMS implementation in SNFs.

**Methods:** A qualitative study of AMS implementation was conducted in Southern Arizona SNFs randomly sampled to represent urban/suburban, border, and rural regions. Semi-structured interviews were conducted with administrators, clinicians, and nonclinical staff within participating facilities. Interview transcripts were analyzed using constant comparative analysis, with both directed and emergent coding, facilitated by NVivo 12 software.

**Findings:** From 04/13/2019 through 12/13/2019, 57 interviews were conducted with 9 administrators, 38 clinical providers, and 10 nonclinical staff across 6 urban/suburban, 2 border, and 2 rural facilities. Analysis identified two thematic categories: "influencer themes," which describe specific barriers and facilitators to AMS implementation, and "system themes," which characterize SNFs as complex adaptive systems shaped by interacting staff roles, care transition challenges, and differing perceptions of AMS practices within the same facility. Key facilitators included effective internal communication, ongoing AMS education, and clinician AMS champions. Primary barriers included poor interfacility communication during care transitions, limited access to diagnostic resources, enculturated prescribing norms, and tension between immediate infection control priorities and long-term AMS goals.

**Conclusions:** Findings suggest that AMS implementation in Arizona SNFs is best understood as a systems-level process emerging from interactions among staff roles, organizational workflows, and care transitions, rather than solely from individual prescribing decisions. Recognizing SNFs as complex adaptive systems highlights the importance of communication structures, local champions, and feedback mechanisms. It underscores the need for coordination strategies within and across SNFs to sustain AMS interventions.

## Introduction

The non-judicious use of antibiotics in healthcare settings drives antibiotic resistance, which is a significant public health threat[1]. Skilled nursing facilities (SNFs) face unique challenges in antimicrobial stewardship (AMS) due to their patient populations and care delivery systems within congregate living settings. Over 60% of long-term care residents are aged 65 or older, making them particularly susceptible to infections due to weakened immunity, multiple comorbidities, and physical limitations[2]. This vulnerability drives high antibiotic use as healthcare providers seek to treat suspected infections expeditiously to prevent individual illnesses and within-facility transmission. Over 70% of SNF residents receive one or more antibiotic courses annually, with 40-75% deemed inappropriate or non-compliant with clinical guidelines[3, 4]. The consequences of inappropriate prescribing include the proliferation of multidrug-resistant organisms[3, 5-7], and heighten residents’ risk of adverse drug reactions, polypharmacy, and decreased antibiotic efficacy due to altered pharmacokinetics[8]. A systematic review of the literature shows that AMS interventions in SNFs are associated with improved health outcomes among residents[9], underscoring the need for effective dissemination of stewardship programs.

Several factors complicate antibiotic prescribing in SNFs. First, long-term care residents are frequently prescribed antibiotics for suspected infection with incomplete clinical or microbiological evidence of infection[10]. Secondly, asymptomatic bacteriuria is common in older adults so that antibiotics may be prescribed in the absence of clinical signs or symptoms of infection[11]. Collectively, these SNF prescribing practices may be characterized as defensive medicine. For instance, a large study in Europe found that over 1 in 4 antibiotics prescribed for urinary tract infections (UTIs) were prophylactic[12]. Similarly, in U.S. nursing homes, a quarter of antibiotics were prescribed for UTI prophylaxis, with a planned duration of over 100 days, compared with the 7 days normally used for treatment, reflecting the use of standing orders for many patients[10]. In addition, nursing homes typically have 2-10 prescribers, and variation in practice patterns may lead to increased antibiotic initiations and decreased accountability for overall antibiotic use[13]. In sum, prescribing decisions in skilled nursing facilities are influenced by patient-specific factors, caregiver expectations, the clinical setting, the healthcare system, and the prescribing culture[14].

Antimicrobial stewardship (AMS) aims to reduce adverse patient outcomes and prevent resistance through judicious, evidence-based prescribing[3]. AMS involves coordinated strategies to optimize antibiotic use, including selecting the appropriate drug, dose, duration, and route of administration to slow the development of resistance[15]. Prior qualitative research suggests that nursing staff and medical providers generally support antimicrobial stewardship goals but face substantial challenges related to diagnostic uncertainty, communication, and competing clinical priorities in nursing home settings[16]. Moreover, the COVID-19 pandemic has further complicated AMS efforts[6, 17], underscoring the ongoing need to understand implementation barriers.

Antimicrobial stewardship efforts in skilled nursing facilities began emerging before the 2015 CDC Core Elements for Nursing Homes and expanded as national guidance and CMS regulatory requirements increased expectations for stewardship programs. However, implementation approaches continued to vary widely across facilities. A 2016 survey of Arizona’s long-term care facilities revealed that only 18% of the 56 facilities that responded met the Centers for Disease Control and Prevention’s seven core elements of stewardship[18]: leadership commitment, accountability, pharmacy expertise, action, tracking, reporting, and education. One thing to note is that there was a very low response, suggesting that actual rates of implementation were much lower. In particular, the mechanisms underlying SNF antibiotic prescribing practices remain poorly understood.

Despite increasing expectations for antimicrobial stewardship, less is known about how stewardship programs are implemented within the complex organizational environments of skilled nursing facilities and how contextual factors shape implementation. We therefore aimed to identify factors that facilitate and hinder AMS implementation in SNFs. Employing a systematic grounded theory approach, this study aimed to understand the process of AMS implementation in Arizona’s urban/suburban, rural, and border SNFs. Our qualitative approach, guided by a sensitizing framework of barriers and facilitators, allowed us to capture the complexity of AMS implementation in varied settings and develop a substantive theory of how these facilities operate as complex adaptive systems, with broad implications for SNFs more broadly[19].

## Methods

### Study design and setting

We used a systematic grounded theory approach to explore and generate a theory of AMS implementation in SNFs[20]. This methodology enabled the emergence of novel theoretical explanations grounded in empirical data, while acknowledging the researchers’ theoretical sensitivity to the topic.

Our team of public health-oriented researchers conducted this study with Arizona-based SNFs between April 13, 2019, and December 13, 2019. The SNFs were selected using publicly available provider information from the Centers for Medicare & Medicaid Services (CMS) database [21]. Facilities were randomized within three strata: urban and suburban facilities along the Phoenix-Tucson corridor, those in the US-Mexico border region (defined as within 100 km of the border), and facilities designated as rural by the Arizona Rural Health Commission. Activities underwent full review by the University of Arizona IRB. Approval #: 1809957428.

### Recruitment and data collection

The study was conducted at 10 facilities in Arizona. At each facility, the administrator served as the entry point: All administrators approved recruitment before scheduling interviews, and we coordinated with them to connect with staff deemed appropriate for interviews. All staff who agreed to participate in the study at each of the 10 facilities were scheduled for interviews on the same day. If the medical directors or other administrators were not on site, we scheduled Zoom interviews with them; the primary data collection method was in-person. Participants provided written informed consent before beginning in-person interviews and oral consent before beginning Zoom interviews. We used maximum heterogeneity sampling, which seeks to maximize the diversity of the respondents’ roles.

Once a facility’s administrator agreed to participate, they signed an attestation form to allow researchers to come on-site to conduct interviews. Researchers set a date for in-person interviews of administrators, clinical care providers, and non-clinical staff. In some cases, medical directors were not on site, so a Zoom interview was scheduled for a date following the site visit. Consent was obtained before the interview: participants signed a consent form for in-person interviews, while audio consent was recorded for phone interviews. All interviews were audio-recorded, transcribed verbatim, checked against the recordings for accuracy, and anonymized. The respondents were: administrators, including executive directors (ED), medical directors (MD), directors of nursing (DON), or infection preventionists (IP); clinical care providers, including registered nurses (RN) and certified nursing assistants (CNA); and non-clinical care providers, including housekeeping and environmental services staff.

A semi-structured interview guide was originally created based on a literature review and the CDC’s AMS framework, which served as sensitizing concepts to guide early data collection around the study’s scope of uncovering facilitators and barriers. Questions were designed to assess antibiotic use in SNFs and included scenarios related to sepsis and UTI management. Semi-structured interview scripts were designed to capture data on work responsibilities, knowledge, attitudes, and perceptions of antibiotic resistance and use, and AMS implementation at their facility. Each interview lasted 60-90 minutes. The guide contained open-ended questions designed to elicit respondents’ opinions about implementing AMS interventions, their experiences managing or interacting with patients regarding antibiotics, and their perceptions of the barriers and facilitators to antibiotic stewardship in the SNF setting. However, some sections were different for non-clinical staff.

### Data analysis

Transcribed interviews were imported into NVivo 12 (QSR International) for analysis. We used constant comparative analysis, incorporating both directed and axial coding to inductively identify themes while allowing a priori themes from the research literature to inform the creation of our codebook[22]. Primary coding was done by three reviewers who independently reviewed six interviews, and interrater reliability was assessed to ensure consistency. The coding process was repeated twice until a high interrater reliability was achieved (κ=0.8)[23]. The finalized, standardized codebook was then used by two reviewers to code all transcripts independently, with a third reviewer resolving discrepancies as necessary. Coders periodically revisited the codes to maintain consistency. Finally, the coded data were examined to identify overarching themes, and they were further queried to explore relationships and patterns across the dataset, culminating in the emergence of a substantive theory.

## Results

A total of 57 semi-structured interviews were conducted with staff from 10 Arizona SNFs (Table 1). Of the 10 participating SNFs, six were urban/suburban, two were border, and two were rural as defined by the Federal Office of Rural Health Policy (FORHP) based on 2010 census data[24]. The largest category of respondents (56.1%, n = 32) were nurses or nurse aides —including IPs, RNs, DONs, and bedside caregivers—and 71.9% (n = 41) had more than 5 years of experience.

**Table 1.** Characteristics of interview respondents at 10 skilled nursing facilities in southern AZ.

| *Recruitment categories* | *Respondent's role* | *Frequency n (%)* | *Years of experience* |  |  | *Hours spent at the facility per week* |  |  |
| --- | --- | --- | --- | --- | --- | --- | --- | --- |
|  |  |  | *<5* | *5-10* | *>10* | *<20* | *20-40* | *>40* |
| *Administrators* | Administration <sup>a</sup> | 8 (14.0) | 5 | 1 | 3 | 0 | 3 | 6 |
|  | Director of Nursing | 9 (15.8) | 1 | 7 | 1 | 1 | 1 | 7 |
| *Clinical care givers* | Bedside Caregivers <sup>b</sup> | 10 (17.5) | 3 | 1 | 6 | 0 | 8 | 2 |
|  | Infection Preventionist | 6 (10.5) | 0 | 5 | 1 | 1 | 5 | 0 |
|  | Medical Doctor | 6 (10.5) | 0 | 1 | 5 | 6 | 0 | 0 |
|  | Registered Nurse | 7 (12.3) | 2 | 3 | 2 | 2 | 5 | 0 |
| *Non-clinical staff* | Housekeeper | 10 (17.5) | 5 | 3 | 2 | 0 | 10 | 0 |
|  | Environmental Services Director | 1 (1.8) | 1 | 0 | 0 | 0 | 0 | 1 |
|  | Total | 57 (100) | 16 (28.1) | 21 (36.8) | 20 (35.1) | 10 (17.5) | 32 (56.1) | 15 (26.4) |

<sup>a</sup> Includes Administrator, Case Manager, and Executive Director
<sup>b</sup> Includes both CNAs and LPNs

Our analysis of transcribed interviews identified two complementary perspectives on AMS implementation. ‘System themes’ (Table 2) describe organizational structures, professional roles, and care transitions that shape the context for AMS. ‘Influencer themes’ (Table 3) identify specific barriers and facilitators operating within these systems.

**Table 2.**
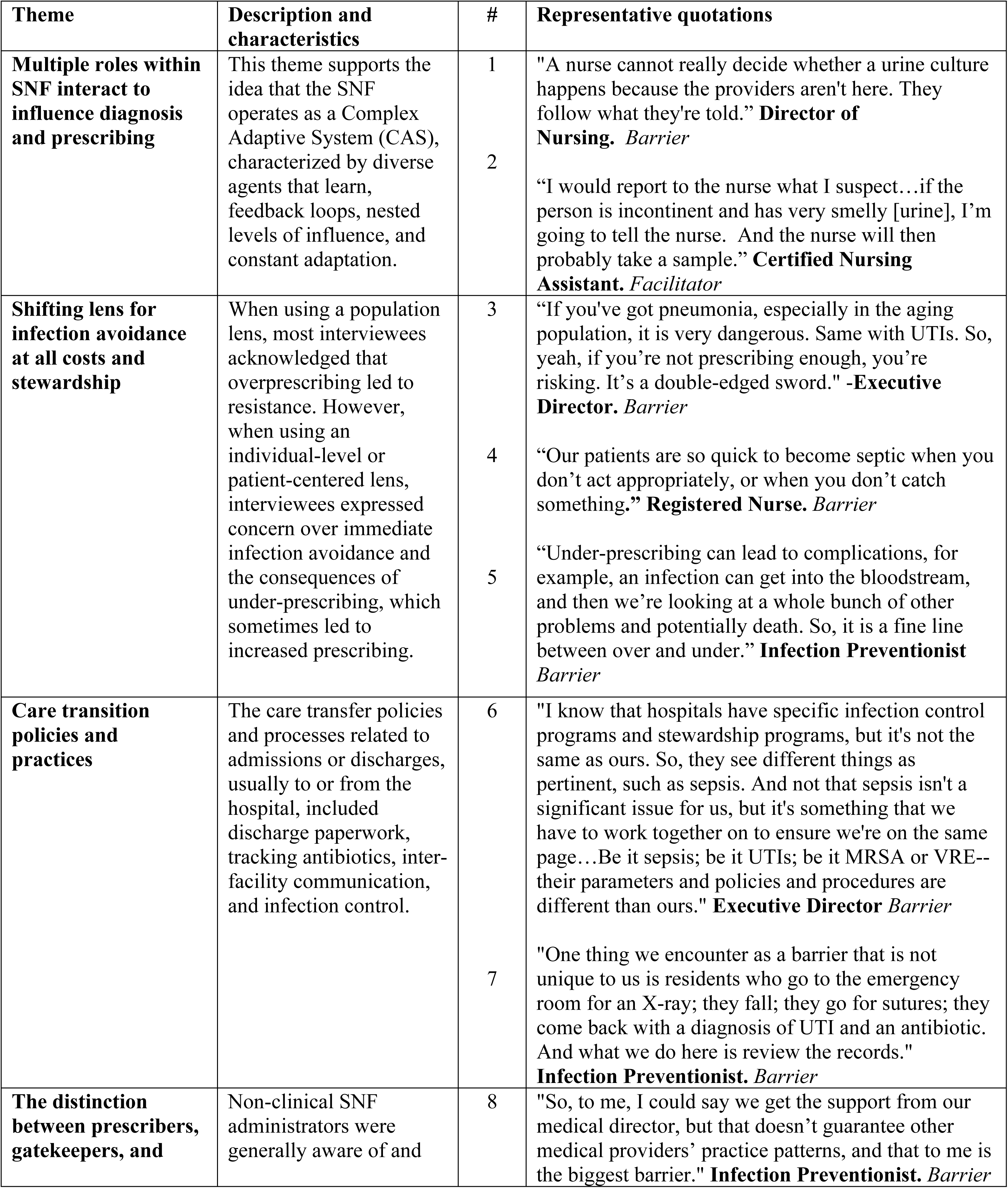
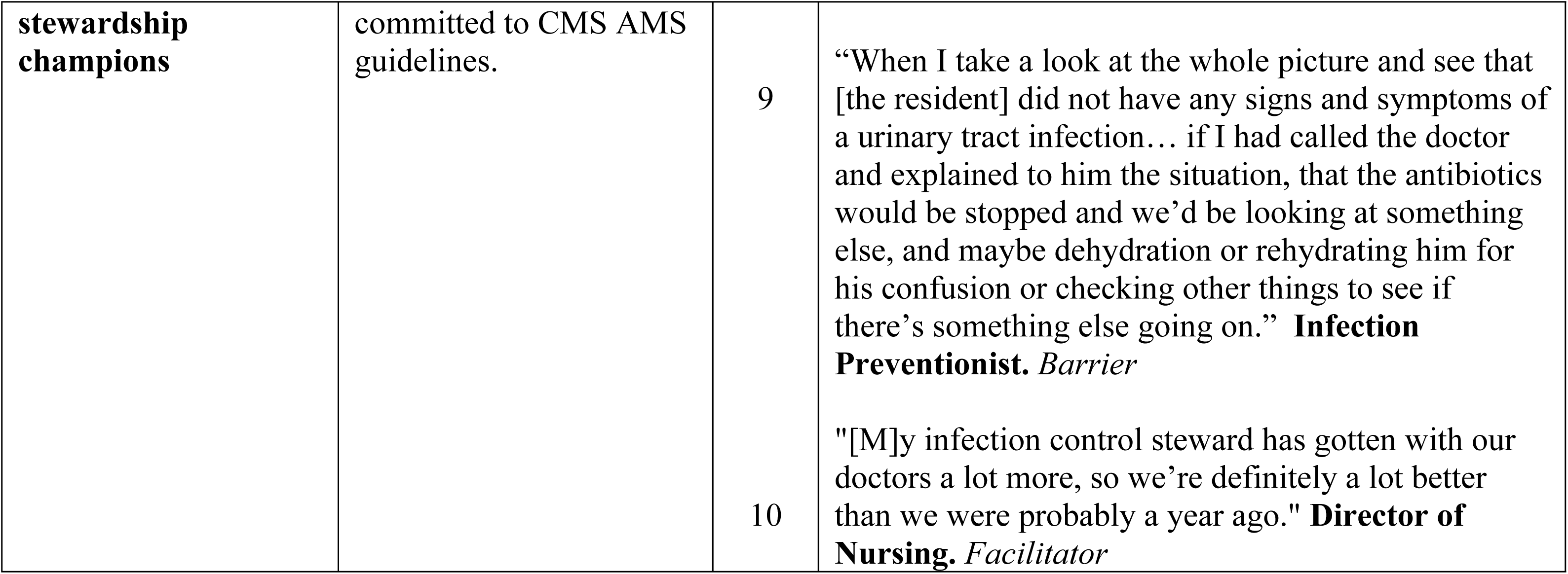
System-level themes reflecting organizational dynamics, roles, and relational structures that influence antimicrobial stewardship with representative quotations.

**Table 3.**
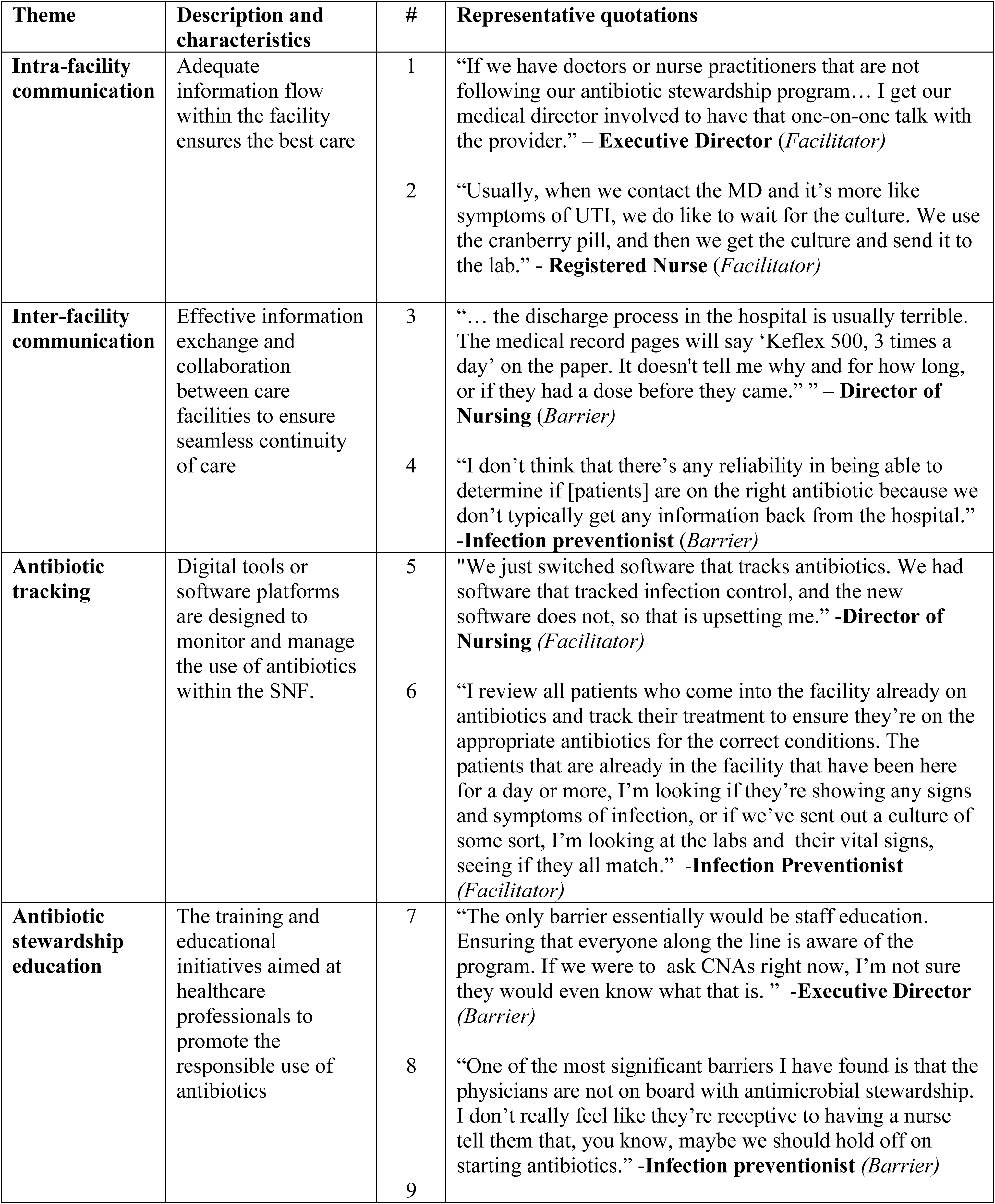
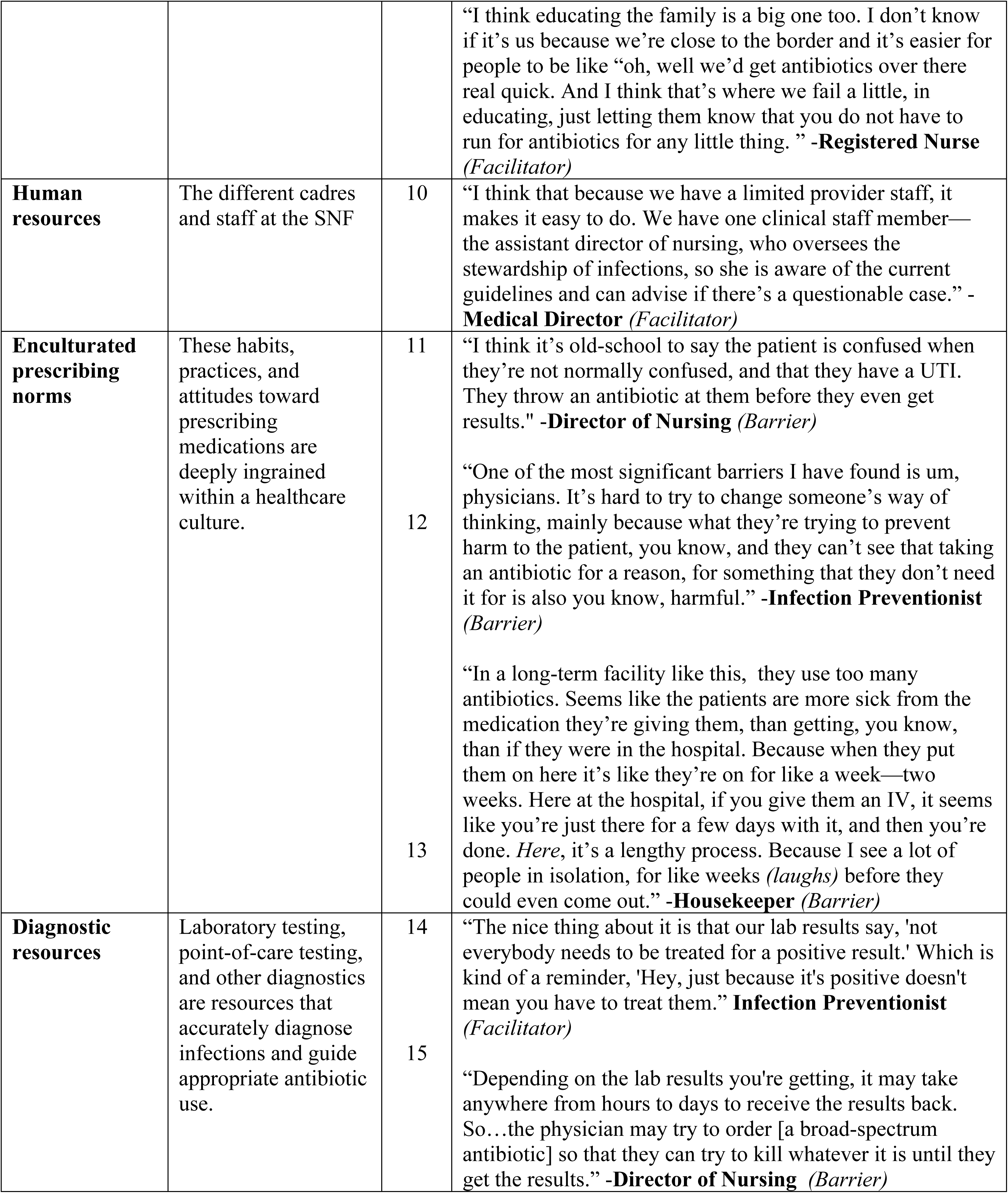
Influencer themes that were characterized as both barriers or facilitators to antimicrobial stewardship in skilled nursing facilities with representative quotes.

### System themes

Systems themes highlight the organizational dynamics, individual roles, and relationships within the complex environment that, together, shape outcomes (Figure 1). There were four Systems themes identified:

**Figure 1.**
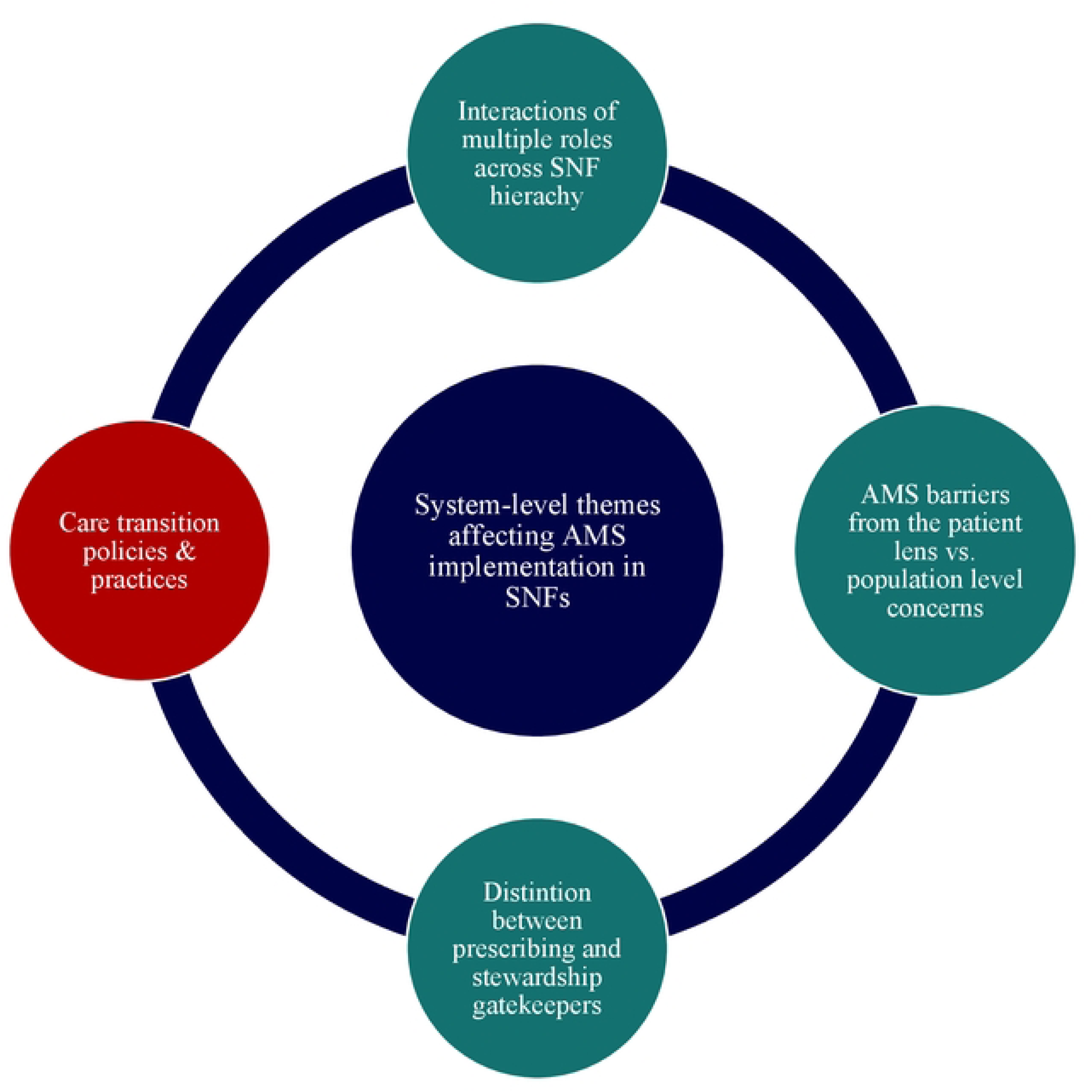
System-level themes impacting successful antimicrobial stewardship implementation with green circles aligning with themes explicitly addressed in the Centers for Disease Control and Prevention (CDC) Core Elements for Antimicrobial Stewardship in Long-term Care, and the red circle indicating a theme not addressed in the CDC core elements.

#### Interactions among multiple roles within the SNF

In addition to housekeepers, all roles (directors, MDs, IPs, DONs, RNs, CNAs, and patient families) directly influenced the decision to order a urine culture or prescribe antibiotics. Aside from IPs (who consistently facilitate AMS), the influence could be positive or negative (Table 2, Quotes 1 & 2). Front-line nurses played a vital role in rapid response, identifying early clinical indicators – such as smelly urine – to initiate culture requests because prescribers were not always on site.

#### Shifting lens for infection prevention and stewardship

The respondents acknowledged how a population-level versus individual-level lens influenced values and practice norms around antibiotic prescribing. Clinicians and administrators were highly attuned to the risk of *not* prescribing, potentially missing the treatment of a real infection in the individual patient. They expressed concerns about the potential repercussions of under-prescribing and acknowledged that worst-case-scenario fears occasionally led to more prescriptions (Table 2, Quotes 3-5). As a result, AMS best practices were often perceived as conflicting with patient safety, highlighting tensions between individual-level patient care concerns, population-level public health concerns about antimicrobial resistance, and existing AMS protocols.

#### Care transition policies and practices

Respondents highlighted the importance of care transitions, care transfer policies and processes related to admissions or discharges, usually to or from the hospital, including discharge paperwork, antibiotic tracking, inter-facility communication, and infection control. The hospital played a key role in the SNF’s ability to track antibiotics and manage patient care. Positive feedback loops with the hospital (e.g., clear documentation of antibiotic prescribing) simultaneously helped the SNF to support patients and AMS. Negative feedback loops (e.g., unclear duration and/or indication for the antibiotic) inhibited patient care and AMS support (Table 2, Quotes 6-7). These inter-facility processes underscore that, as open systems, SNFs interact within larger healthcare systems whose policies and procedures shape SNF practices.

#### The distinction between prescribers and stewardship champions

Non-clinical SNF administrators were generally aware of and committed to CMS guidelines for AMS. AMS compliance often fell to administrators, IPs, and DONs; however, prescribers were ultimately responsible for determining the prescribing practices at the facility. These prescribers acted as AMS barriers when they did not follow, or incorrectly implemented, stewardship practices (Table 2, Quotes 8-9). Conversely, prescribers were AMS facilitators when they actively championed stewardship (Table 2, Quote 10). Across all roles, interviewees perceived prescribers as ultimately responsible for AMS. When prescribers had positive relationships with IPs or DONs, or when prescribers themselves were stewardship champions, positive feedback loops promoted stewardship. When relationships were more adversarial or the prescriber did not champion AMS, barriers emerged.

### Influencer themes

The influencer themes highlighted barriers and facilitators to effective AMS implementation, including communication between and within facilities, AMS education and tracking, and human and diagnostic resources. There were seven influencer themes identified.

#### Intra-facility Communication

Effective communication among care providers at different levels within SNFs was crucial for AMS, and respondents in the administration category serve as the go-between to ensure it. When information moved freely and often laterally and hierarchically among care providers, communication acted as an AMS facilitator (example, during shift changes, asynchronously as test results became available, when patient status changed, etc.). When the flow of information was restricted (example, by rigid hierarchy, omission during shift changes, etc.), communication acted as an AMS barrier *(Table 3, Quotes 1-2)*.

#### Inter-facility Communication

The respondents in the clinical care and administration categories mentioned that the gap in the management of residents was in communication with hospitals, particularly regarding information flow on prescriptions, which differs from the structural components (e.g., policies and established systems) that govern care transfer. For example, when patients were discharged from hospitals with antibiotic prescriptions, critical details such as the intended duration of therapy or the clinical rationale were often missing from transfer records *(Table 3, Quote #3).* Moreover, SNFs frequently did not receive follow-up information from hospitals to clarify or justify these prescriptions *(Table 3, Quote #4)*. This breakdown gap posed a significant barrier to AMS, as coordinated care depends not only on structural systems but also on the reliable exchange of accurate, actionable information across settings.

#### Antibiotic Tracking Systems

Antibiotic tracking systems were a critical component of AMS in SNFs. One respondent reported that although their facility has an electronic tracking system, they were still getting used to new software that will aid tracking antibiotic use *(Table 3, Quote #5)*. Manual tracking was regularly also conducted to support AMS efforts *(Table 3, Quote #6).* Other respondents highlighted the presence of quality assurance systems that analyze internal infections and share compliance information monthly, serving as a facilitator to the AMS.

#### AMS Education

Consistent education on prescribing protocols and best practices that engaged staff across multiple job categories was frequently cited as an AMS facilitator. The respondents noted that one of the gaps in embracing AMS was that everyone involved in patient care, including doctors and certified nursing assistants, needed AMS education *(Table 3, Quotes #7-8)*.According to many respondents, aligning the team’s antibiotic beliefs would strengthen the AMS program and coordinate efforts. The respondents also mentioned efforts to educate patients, families, and caregivers about AMS to address the demands for antibiotic prescription *(Table 3, Quote #9)*.

#### Human Resources

Respondents reported that SNF’s robust human resources affected AMS. Smaller teams might simplify decision-making and reduce redundancy, guaranteeing that protocols are followed *(Table 3, Quote #10)*. Respondents stated that human resource constraints could increase accountability—they forced everyone to participate in training and reduced the number of nursing supervisors.

#### Enculturated Prescribing Norms

Enculturated prescription norms had a substantial influence on AMS practices. Some healthcare providers administered antibiotics before test results were available, adhering to the conventional wisdom that once a patient is confused, that indicates an infection and necessitates an antibiotic *(Table 3, Quote #11)*. Respondents described this as a still-common practice among many physicians that contributed to overprescribing, sometimes making patients sicker (Table 3, Quotes *#12-13)*.

#### Diagnostic Resources

The availability and utilization of diagnostic resources were crucial for supporting AMS assessment and management. The turnaround time for laboratory results directly influenced AMS efforts and was also critical to resident well-being (Table 3, Quote#14). In some cases, broad-spectrum antibiotics were prescribed before lab results were available, not to undermine diagnostic testing but to address concerns about infection progression, and to kill whatever it is before they get the results *(Table 3, Quote #15)*.

## Discussion

In this study, we found that the system themes portrayed SNFs as a diverse group of agents that learn, exhibit feedback loops, and undergo constant adaptation. Individuals in IP roles often had additional responsibilities as both nurses and IPs. These roles were more likely to identify as AMS facilitators, see themselves as educators, and effectively respond to AMS concerns. Effective AMS programming depends on communication between prescribers, DONs, and IPs. As staff worked to balance patient safety and prescribing concerns, the perceived conflict between infection prevention and AMS was a significant obstacle. Furthermore, attempts to ensure AMS programming were hampered by communication gaps during the care transition, underscoring the importance of hospital involvement in AMS.

While staffing constraints hindered conversations about antibiotic usage, effective intra-facility communication and sufficient staffing enhanced decision-making and support AMS. Smaller teams encouraged accountability, whereas antibiotic-tracking systems and consistent AMS training for all staff improve adherence. Inter-facility communication was problematic, particularly when exchanging antibiotic-related information between hospitals and SNFs. Enculturated prescribing practices hindered AMS. While delayed test results may lead to early antibiotic prescriptions, prompt diagnostic tools were crucial for AMS.

### Facilitators

Previous research has shown that nurses are consistently present in healthcare facilities, are often perceived as AMS gatekeepers, and are more likely to influence prescribing decisions because they usually double as IPs[25-27]. Our findings align with this, indicating that nurses had multiple roles and were more likely to champion AMS initiatives. Respondents emphasized the importance of communication amongst colleagues, particularly when addressing non-compliance with AMS protocols, reinforcing previous findings that effective communication is critical to the success of AMS programs[28]. Regular and ongoing education for all staff was identified as one of the key facilitators for AMS in previous studies[25, 27-29], echoed by our participants, who noted that increased staff education would enhance AMS adoption within facilities. Additionally, respondents highlighted the value of digital tools to monitor antibiotic use, aligning with recommendations from prior research in nursing home settings[27].

### Barriers

Fear of missing an infection, the unpredictability of patient populations, and concerns that infections could escalate out of control have been identified in the literature as key barriers to the effective implementation of AMS[16, 26, 28, 29]. In our study, the tension between infection control priorities and AMS goals emerged as a significant barrier. Respondents described a clear distinction between AMS gatekeepers and prescribers, with nurses frequently identifying non-compliance and taking AMS champion roles, an issue noted in prior research[25, 26].

External factors, such as poor communication during care transitions and emergency department visits, have been shown to disrupt and undermine AMS endeavors[27-30]. These findings were echoed in our study, which highlighted gaps in care transition practices as significant obstacles.

Additionally, a strong reliance on antibiotics, often driven by prescribers’ perceptions and entrenched prescribing norms sometimes referred to as ‘old school’ practices, was noted by our respondents and has been similarly reported in earlier studies. Finally, limited access to diagnostic resources was frequently cited as a barrier to AMS, consistent with other research indicating that a lack of timely diagnostic information hampers appropriate antibiotic decision-making[26, 28, 29].

### Limitations and strengths

This study has limitations. First, we conducted interviews in 2019, before the COVID-19 pandemic disrupted healthcare delivery and AMS programs nationwide. However, AMS in SNFs remains a persistent challenge[31-33], with increased urgency due to rising rates of carbapenem-resistant Enterobacteriaceae (CRE) and *Candida auris* in Arizona and nationwide. The barriers and facilitators we identified—organizational dynamics, care transitions, and professional role tensions—remain central to AMS implementation regardless of pandemic context.

Second, we sampled 10 of 145 Arizona SNFs, which may limit generalizability. However, our purposeful sampling across urban, suburban, rural, and border facilities captured diverse organizational contexts and maximized heterogeneity in facility characteristics and staff perspectives. This diversity enhances the transferability of findings to similar SNF settings.

A key strength of this study is its qualitative approach across multiple staff roles and facility types, revealing systems-level dynamics that quantitative studies cannot capture. The CAS framework provides a novel lens for understanding how organizational structures, professional relationships, and care transitions shape AMS implementation.

## Conclusion

Our findings demonstrate that effective AMS in SNFs requires a systems approach addressing both intra-facility dynamics (professional roles, communication hierarchies) and inter-facility coordination (care transitions with hospitals). The CAS framework reveals why piecemeal interventions fail: SNFs operate within complex healthcare systems where hospital policies, prescriber practices, and organizational cultures interact to shape stewardship outcomes. Successfully implementing CDC’s Core Elements requires not only internal commitment but also strengthened communication protocols during care transitions, which remain under-addressed in current guidance.

## Data Availability

This is qualitative data that can be shared on request.

## Acknowledgement

We acknowledge Laura Shriver and Ferris Ramadan, who participated in the data collection, and the staff from the 10 SNFs that participated.

